# PERFORMANCE OF GOOGLE TRENDS FOR EARLY DETECTION OF DENGUE INFECTION EPIDEMICS IN JAKARTA AND YOGYAKARTA

**DOI:** 10.1101/2020.02.19.20024323

**Authors:** Atina Husnayain, Setyarini Hestu Lestari, Siti Nadia Tarmizi, Anis Fuad

**Affiliations:** Department of Biostatistics, Epidemiology and Population Health, Faculty of Medicine, Public Health and Nursing, Universitas Gadjah Mada, 55284, Yogyakarta, Indonesia; Center for Health Policy and Management, Faculty of Medicine, Public Health and Nursing, Universitas Gadjah Mada, 55284, Yogyakarta, Indonesia; Yogyakarta Provincial Health Office, 55244, Yogyakarta, Indonesia; Directorate of Vector-borne and Zoonotic Diseases, Directorate General of Disease Control and Prevention, Indonesian Ministry of Health, 10560, Jakarta, Indonesia

**Keywords:** dengue, digital epidemiology, Google Trends, Indonesia

## Abstract

**Background:** Early detection of disease outbreak is among the most critical role of the sub-national authorities as mandated by the health decentralization policy. Given the continuous growth of Internet penetration and dependencies of the society on the digital ecosystem, it is essential to investigate the potential innovations to improve the existing surveillance system using digital epidemiology. Several studies, including in Indonesia, have assessed the roles of Google Trends (GT) to improve dengue surveillance systems. However, they were mostly located in specific areas or national level only. No reports are available to compare the performance of GT for early detection of dengue outbreak among high burdened provinces.

**Aims:** This study aimed to examine the correlation between GT data on dengue-related query terms with the official dengue surveillance reports in Jakarta and Yogyakarta Province.

**Methods:** Relative Search Volume of GT data for dengue were collected from the area of Jakarta and Yogyakarta between 2012 to 2016. Those data were compared with the official dengue reports from the Indonesian Ministry of Health using Pearson’s correlation and Time-lag correlation, performed with Stata version 13.

**Results:** GT data are positively correlated with the routine surveillance report in Jakarta (*r* = 0.723, *p*-value= 0.000) and Yogyakarta Province (*r* = 0.715, *p*-value= 0.000). In Jakarta, search term of ‘DBD’ demonstrated a very strong correlation for lag-1 (*r* =0.828, *p*-value= 0.000). This finding indicates that GT data could possibly detect the dengue outbreak a month earlier, especially in Jakarta. Hence, GT data can be used to monitor disease dynamics and improve the public awareness of a potential outbreak in near-real-time.

**Conclusion:** GT data were positively correlated with the routine surveillance report in Jakarta and Yogyakarta Province. Early warning system utilizing GT data is potentially more accurate in Jakarta than in Yogyakarta. We assume that it is related with the larger population as well as the Internet use activities that drives the higher volume of Google search on dengue in Jakarta compared to Yogyakarta. Further studies involving other digital data sources, for example, Twitter, online news, and administrative data from the national health insurance are essential to strengthen the current surveillance system with the new digital epidemiology approach.

## INTRODUCTION

Dengue virus infection is one of the major public health problems in Indonesia. Since the first report in 1968, dengue cases have infected all regions and repeatedly caused outbreaks with notable disabilities and casualties. At the global level, dengue is among the diseases with the highest increase of incidence rates, against the global trend of lessening communicable diseases (Karyanti *et al*., 2014; Caballero-Anthony *et al*., 2015; Stanaway *et al*., 2016; Katzelnick, Coloma and Harris, 2017).

Currently, no provinces in Indonesia are free from the disease. In five decades, Java island, the home of about 60% of Indonesians was the most significant contributor to dengue cases (Harapan *et al*., 2019). In 2017, Jakarta and Yogyakarta Provinces were among the most dengue-burden areas, with the incidence of 32.29 and 43.65 per 100,000 population, respectively. Dengue infection incidences in these two smallest provinces in terms of the areas were significantly higher than the national incidence, which was about 22.55 per 100,000 population (Indonesian Ministry of Health, 2018).

Given the consideration as a priority disease that repeatedly causes outbreaks, a robust surveillance system is needed. Unfortunately, the existing dengue surveillance system has suffered from several issues, namely underreporting, limited timelines, inadequate completeness, and other concerns (Runge-Ranzinger *et al*., 2014; Das *et al*., 2017). In Bandung regency, the timeliness of dengue surveillance report varies from days to months (Adrizain, Setiabudi and Chairulfatah, 2018). Therefore, innovations to strengthen the surveillance system, especially in detecting potential dengue outbreaks in an earlier and more proactive manner become essential.

Digitalization and revolution in communication have changed human behavior and culture. In this regard, the World Health Organization published recommendations on digital interventions to strengthen the global health systems (World Health Organization, 2019). Thus, the science of epidemiology, which has long studied the risks and distribution of health-related states and events in the population, began to recognize a new approach, namely digital epidemiology. Various digital data sources and traces from multiple platforms that are used routinely by the community are potential sources to understand the dynamics of health-related states and events in the population (Salathé *et al*., 2012; Salathé, 2018).

With billions of users googling for information every day, the search traces left by users become highly valuable. Google Trends (GT) is a freely accessible platform provided by Google Inc., allowing to explore the patterns of particular “keywords” entered by Internet users while searching for information. Aggregated data of Internet search queries can be sorted based on geographical location and time. The scale of relative search volume (RSV) is from 0 to 100, where 100 is the highest search rate of the term in a given period (Rogers, 2016).

GT publications in healthcare increased significantly in a variety of topic domains using various methods. For the public health domain, GT has increasingly received attention as a potential data source to complement and even improve the existing disease surveillance programs (Nuti *et al*., 2014). Several GT studies on Zika, influenza, chikungunya, and dengue infection have produced promising results (Kang *et al*., 2013; Teng *et al*., 2017; Mahroum *et al*., 2018; Morsy *et al*., 2018).

In our previous study, GT showed potential as an early warning tool for dengue infection surveillance systems at the national level (Husnayain, Fuad and Lazuardi, 2019). A prediction model of dengue infection dynamics for Surabaya has been developed based on reports from a single referral hospital (Anggraeni and Aristiani, 2017). In the United States, the capability of GT to monitor pertussis shows different results between states (Arehart, David and Dukic, 2019). To the best of our knowledge, studies comparing the performance of GT between sub-national authorities in Indonesia do not exist yet. Due to the decentralization policy and the critical role of local governments to declare epidemic status, such study is essential.

In this regard, we aimed to study the performance of GT in two unique provinces in Java, namely Jakarta and Yogyakarta Province. They are selected based on two reasons: high dengue-burden areas as well as high Internet penetration. At the national level, 91.45% of the population aged over five years living in urban areas have accessed the Internet during the last three months. In Jakarta and Yogyakarta, this number attained 92.87% and 94.40%, respectively (BPS-Statistics-Indonesia, 2017).

## METHODS

This study is part of the infodemiology research project in the Department of Biostatistics, Epidemiology and Population Health, Faculty of Medicine, Public Health and Nursing, Universitas Gadjah Mada that was begun in 2017. In collaboration with the Ministry of Health, we have conducted GT studies in the field of vector-borne and zoonotic diseases, including dengue and leptospirosis.

We applied a similar method as the previous study for data collection and analysis. RSV for particular search terms, namely, “gejala demam berdarah”, “demam berdarah”, and “DBD” were collected from the website of Google Trends (https://trends.google.com/). These were the most significant terms that were correlated with the temporal pattern of dengue cases frequency at the national level. Data at the national level were then filtered geographically by Jakarta and Yogyakarta from 2012 to 2016. Geographic levels of RSV were then aggregated by average search volume per month (Gluskin *et al*., 2014). GT data were then compared with the monthly dengue cases of the two provinces. Provincial data of dengue cases were obtained from the Directorate of Vector-borne and Zoonotic Diseases, Indonesian Ministry of Health. Both GT and the official surveillance data did not contain any protected individual health information; therefore, full review from ethical board was not required. For comparison, dengue cases were normalized with the same interval as GT data, which ranges from 0 to 100.

Pearson’s correlation was used to measure linear relationship between monthly GT data and official surveillance reports. Correlation strengths are categorized from very weak to very strong (Evans, 1996). To measure whether the correlation between GT data and surveillance reports occurring after a specific time interval (lag), we applied a lag time correlation analysis. Statistical analysis was performed using Stata version 13.

## RESULTS AND DISCUSSION

The temporal distribution of dengue cases in Jakarta and Yogyakarta Province from 2012 until 2016 is depicted in Figure 1. Recognized as the most burdened dengue areas in the country, however, their characteristics are indeed different. In terms of cumulative numbers and fluctuation dynamics, dengue infection in Jakarta is more evident than in Yogyakarta. The average number of monthly dengue cases in Jakarta Province is 844.15, while in Yogyakarta Province it is 265.2 cases per month. It is thought to be related to some sociodemographic indicators such as total population, areas, population density, and level of urbanity.

**Figure.1.**
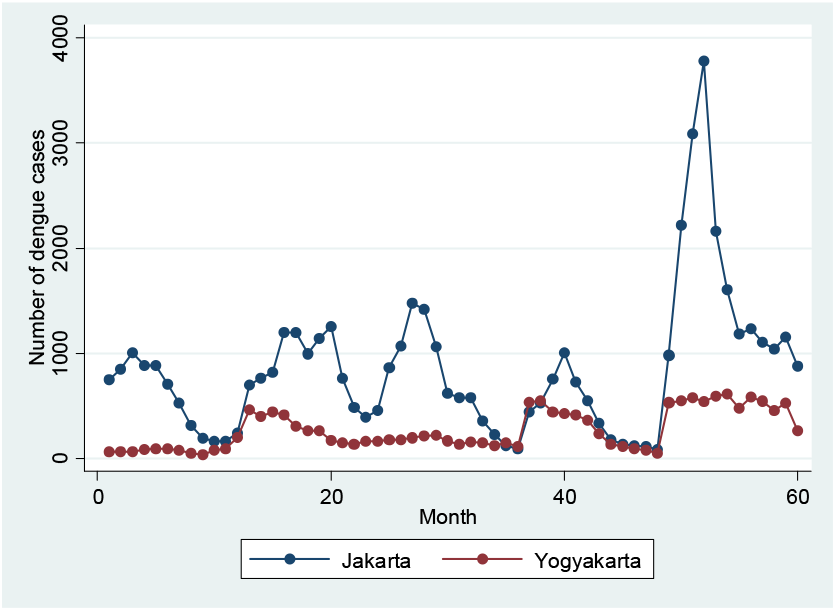
Epidemic curve of monthly dengue cases in Jakarta and Yogyakarta Province (2012-2016)

The epidemic curve that was marked by five peaks confirms the seasonal pattern of the dengue infection. The first four peaks (2012-2015) in Jakarta Province remain stable between 1,004 to 1,476 cases in a month. A far different situation occurred in 2016. The peak of cases per month increased almost four times compared to the previous year. On a national scale, the dengue outbreak occurred in 2016, with an incidence rate of Number of dengue cases 1000 2000 3000 4000 77.96 cases per 100,000 person-years (Harapan *et al*., 2019). In Yogyakarta, the epidemic curve also shown five peaks. However, the peak during the outbreak period in 2016 was relatively similar to the peak in the previous year.

### Correlation Between GT and Official Surveillance Data

Health facilities are among the primary sources for dengue infection surveillance. According to the regulation, hospitals should report any dengue cases to the district health offices within 24 hours after laboratory-confirmed diagnosis (Indonesian Ministry of Health, 2014). However, hospital visits are only the tip of the iceberg of the actual situation. On the other hand, people’s motives for visiting hospitals are influenced by several determinants such as literacy, disease severity, accessibility, referral system, health insurance coverage, and other reasons. Given the incubation period of the dengue virus is about 5.6 days and 95% of cases developed symptoms between 3 to 10 days (Rudolph *et al*., 2014), therefore relying on hospital-based data for dengue surveillance is relatively late to capture the real viral transmission occurring in the community.

GT has the potential to improve the existing surveillance system by providing early signals of occurring disease transmission in the community. One of the underlying reasons is the increase in digital behavior. Googling for information when experiencing any disease symptoms could generate digital traces: the earlier and more frequent searches, the more significant the digital traces for secondary data use.

This study reports a positive correlation between GT and official dengue surveillance reports in Jakarta and Yogyakarta for the following search terms, namely ‘gejala demam berdarah’, ‘demam berdarah’, and ‘DBD’ with various correlation strengths (Table 1).

**Table 1.**
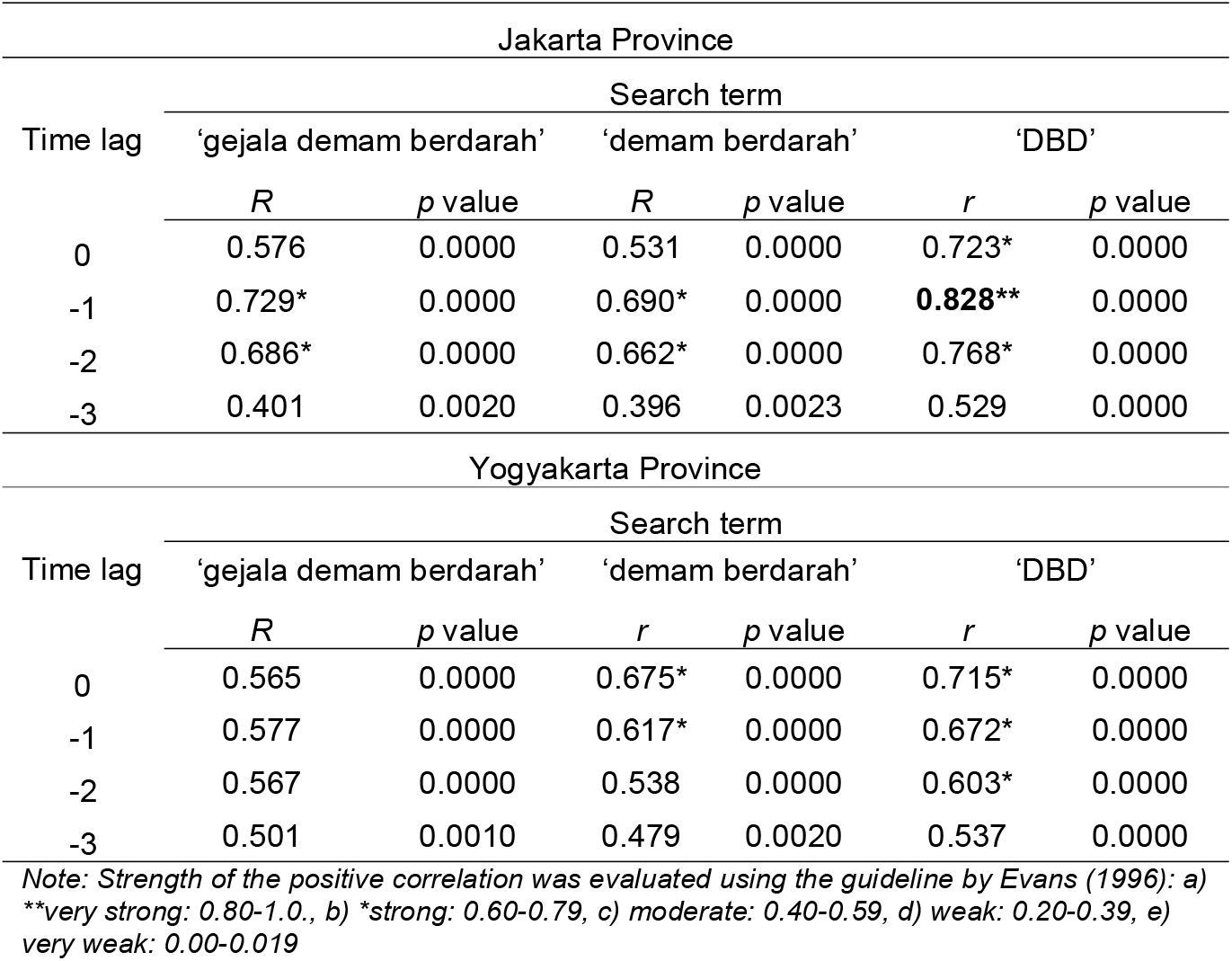
Result of Pearson’s and Time-lag correlation

In Jakarta, all search terms demonstrate a strong positive correlation for lag-1 and lag-2. It means that the rise of monthly dengue cases could be detected within one or two months earlier in GT using the above search terms. Interestingly, the search term of ‘DBD’ has a robust or powerful correlation for lag-1 (*r-value* = 0.828), the highest value compared to other search terms in Jakarta and Yogyakarta for all types of temporal analysis. This search term also shows a strong positive correlation for lag-0.

A different finding is shown in Yogyakarta. Strong positive correlations are exhibited by search terms of ‘demam berdarah’ and ‘DBD’ in lag-0 and lag-1. For lag-2, only search term of ‘DBD’ shows a strong positive, but borderline, correlation.

The above findings confirmed the previous study reporting the variations of GT performance at the sub-national level (Gluskin *et al*., 2014). We suspect that sociodemographic indicators might explain the reason why GT performance in Jakarta is different from Yogyakarta. In 2015, Jakarta was inhabited by 10.2 million population within an area of 664 km2. On the other side, 3.6 million population reside in 3,133 km2 in Yogyakarta. It shows a very contrasting situation in terms of population size and density between the two provinces. From the perspective of urbanity, Jakarta is a metropolitan and capital city. Yogyakarta, on the other hand, is a province consisting of four districts and one municipality with an area stretching from urban, suburban to rural areas (BPS-Statistics-Indonesia, 2017).

A combination of various factors, including the population size, high availability of digital infrastructure, more massive digital behavior, and more intense media coverage, could drive significant “googling practices” and voluminous digital traces in Jakarta than Yogyakarta. Therefore, further studies exploring the connection between those variables with GT performance for early detection of epidemics in greater number of dengue high endemic areas are needed.

The difference of GT performance in the two provinces is seen graphically in Figure 2 and Figure 3. In Jakarta, GT data show a similar pattern with the official surveillance report. The search volume for dengue could be seen to increase one to four months earlier before the rising of dengue cases in epidemic periods (Figure 2). Visually in Figure 3, the pattern shown by GT in Jakarta did not appear in Yogyakarta. Search volume did not rise significantly during epidemic periods in 2013, 2014, and 2015. In 2016, search volume increased rapidly. However, it also decreased rapidly while at the same time dengue cases were still in an epidemic period. The relative search volume in Jakarta was also higher (25.67) compared to Yogyakarta (21.30).

**Figure.2.**
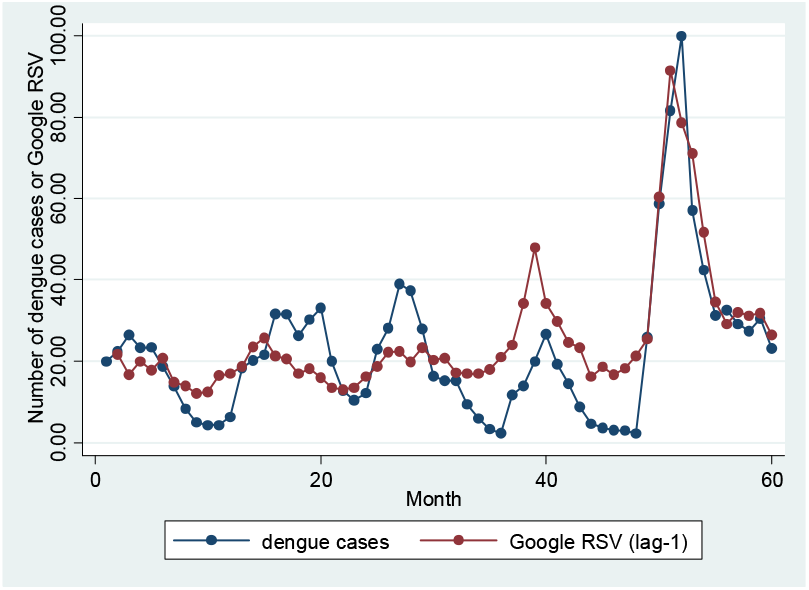
Temporal distribution of monthly dengue cases and relative search volume for dengue (lag-1) in Jakarta Province (2012-2016)

**Figure.3.**
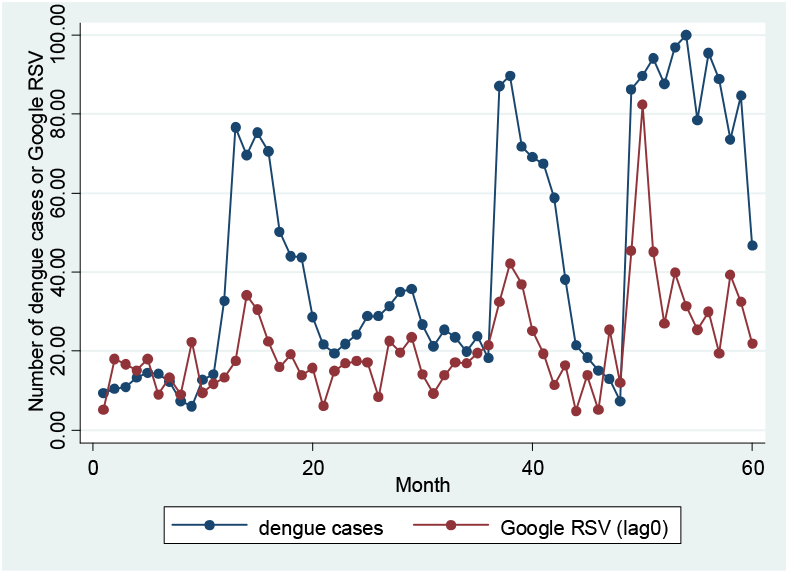
Temporal distribution of monthly dengue cases and relative search volume for dengue (lag-0) in Yogyakarta Province (2012-2016)

### Online Health Information Seeking Behavior

A nationally representative survey by APJII (Indonesian Association of Internet Providers) reported that 51% of Indonesian Internet users search for health information on the internet (APJII, 2017). Another study in California reported quite similar proportions. Based on a survey of 42,087 participants, 53% used the Internet for health purposes (Din *et al*., 2019).

Existing reports on geographical variations and sociodemographic characteristics of online health information seeking in Indonesia are not known. Other studies reported that age (younger than 65 years old), gender (female), level of education (higher level of education), and marital status (married) are primary factors influencing health information-seeking behavior (Beck *et al*.,2014; Nölke *et al*., 2015; Oh and Song, 2017; Din *et al*., 2019).

Further studies to explore motivations for online health information seeking will be essential. In general, reasons for seeking health information could include curiosity, prior knowledge, social expectations, and situational norms (Grasso and Bell, 2015). In the past, patients also sought health information before and after visiting a doctor. However, its intensity has increased since the advent of the Internet and the digital era (McMullan, 2006).

Online health information seeking behavior is also potentially affected by media coverage and mass media exposure. In some cases, it could be the most prominent factor to motivate for information seeking (Alicino *et al*., 2015; Cervellin, Comelli and Lippi, 2017). However, the power of social media for the dissemination and amplification of information could bring fake news and health-related misinformation as well (Wang *et al*., 2019).

Online health information seeking will be a common practice in the future. It is interesting to see how the situation will be changed in the future. The presence of a digital generation, the increasing maturity of digital technology, and equitable access to infrastructure will drive the emergence of more digital innovations in the field of epidemiology.

### Future Use of Google Trends Data

Many studies have reported the use of Google Trends for various public health issues with conflicting results. For certain diseases, a high correlation with official surveillance data occurred, but not for others (Alicino *et al*., 2015; Castro *et al*., 2016; Strauss *et al*., 2016; Cervellin, Comelli and Lippi, 2017; Teng *et al*., 2017). Specifically for dengue, several studies concluded that GT could be used to predict dengue outbreak earlier, with potentially increased sensitivity, and timeliness relative to the traditional surveillance system (Strauss *et al*., 2016). In areas where there is no syndromic surveillance system, GT has the potential as the alternative data source. Indonesia had once implemented a syndromic surveillance system involving several sentinel hospitals, which is now no longer continued (Siswoyo *et al*., 2008).

This research reveals, again, a promising role of Google Trends as a complementary data source for early detection of dengue infection epidemics. This study showed a more favorable finding shows in Jakarta, compared to Yogyakarta. Replicating this study in high endemic provinces is essential given that no provinces in Indonesia are free from dengue. Future research could identify potential areas for developing a dengue early detection system based on GT data. Repeated and ongoing research is essential in line with the government’s policies and programs to expand digital infrastructure development in various regions to reduce digital divides in the country.

Further research is also necessary in certain areas with novel initiatives for dengue vector control. For example, Yogyakarta has started a city-wide release of *Wolbachia*-injected *Aedes aegypti* mosquitoes to reduce the dengue viral transmission (Indriani *et al*., 2018; O’Reilly *et al*., 2019). If this biocontrol intervention is successful, the study of GT in Yogyakarta will likely produce opposite results to the current study.

Innovative research to develop a prediction model by combining GT data with other data sources is also essential. The digital platform, such as Twitter, has been used to predict the spread of dengue within Yogyakarta municipality (Ramadona *et al*., 2016). Environmental, meteorological and climatic data also have the potential to predict the vector dynamics and clinical cases (Carvajal *et al*., 2018; Tosepu *et al*., 2018; Zheng *et al*., 2019).

Another potential data source is the administrative data from the Indonesian Health Security Agency (Ariawan, Sartono and Jaya, 2019). The temporal pattern of dengue cases from 2015-2016 in the Indonesian Health Insurance data sample is similar to the national surveillance report from the Ministry of Health (Fuad, 2019). Indeed, the administrative data was initially collected for claim and reimbursement purposes. However, due to the majority of health facilities, including the primary and referral care, that participate in the universal health coverage system, administrative data has better coverage than the existing dengue surveillance system. Data linkage between these two datasets could generate a more complete picture of the patient journey from the primary to secondary care.

This study also has several limitations. First, we have compared two provinces only, namely Jakarta and Yogyakarta Province. However, based on this study, several recommendations were noted to improve further studies, for example, by involving more dengue-high burden areas in Indonesia. Secondly, this study has validated the GT data with one single source, namely official dengue cases report from the Ministry of Health. With the availability of a national health insurance data sample, further study could also combine with this new dataset. Finally, further studies need also to consider sociodemographic variables to improve the accuracy and explain the variations of GT performance for early detection of dengue epidemics at the sub-national level.

## CONCLUSION

Google Trends data are positively correlated with routine surveillance reports in Jakarta and Yogyakarta Province. An early warning system utilizing Google Trends data is potentially available in Jakarta Province. Further study is needed to replicate this study in other provinces. Studies involving other data sources, for example, digital data sources, environmental data, and administrative data from the national health insurance are necessary to strengthen the existing dengue surveillance system with the digital epidemiology approach.

## Data Availability

Data is available as supplementary file.

## Acknowledgement

We gratefully thank to the Directorate of Vector-Borne and Zoonotic Diseases under the Directorate General of Disease Prevention and Control within the Ministry of Health Republic of Indonesia for providing the official dengue surveillance data from 2012-2016.

